# Infected surfaces as a source of transmissible material in healthcare settings dealing with COVID-19 patients

**DOI:** 10.1101/2021.08.06.21261491

**Authors:** GD Khedkar, Pramod Bajaj, Amol Kalyankar, Rajeshree Deolalikar, Vikram Khilare, Aniket Khedkar, Rahul Bajaj, Chandraprakash Khedkar, Bharathi Prakash, Chaitali Khedkar, Sunil Chavan, P. Jyosthna, Vidya Niranjan, Manju Jilla, Unmesh Takalkar

## Abstract

The disease COVID-19 has turned out to be a tremendous slayer and has had some of the most devastating impacts on human beings ever seen in history. To overcome this major public health crisis, an understanding of the transmission of the virus underlying this disease is of paramount importance. Evidence suggests that the most common route of transmission for the SARS-CoV-2 virus is likely via direct contact in person-to-person encounter with aerosol droplets. However, the possibility of transmission via contact with fomites from surfaces is a possible route of infection as well. Environmental contamination in rooms with COVID-19 patient has been widely observed due to viral shedding from both asymptomatic and symptomatic patients. Also, in hospitals, SARS-CoV-2 is known to survive on various surfaces for extended periods of time. Because repetitive contact cycles can spread the virus from one surface to the other in healthcare settings, here we evaluated contamination on different types of surfaces commonly found in healthcare settings. Also, based on various datasets, we analyzed the importance of various surfaces in transmission modalities. Based on the findings of this study, decontamination of surfaces that frequently are in touch contact throughout all segments of the healthcare system should constitute an important part of the infection control and prevention of COVID-19. We also recommend the selection of a non-reactive disinfectant for hospital monitors, devices, ventilators and computers so that active surface disinfection can be effected without damage to the devices.

## Introduction

The disease COVID 19 and the transmission of the SARS-CoV-2 virus has had one of the most devastating impacts on human beings ever known in history. After two successive waves of SARS-CoV-2 infections in just over one year, we are still debating the role and importance of transmission modes of SARS-CoV-2. This is reflected in part by the only cursory reference given to this issue in many infection control guidelines [1-3]. According to current evidence, SARS-CoV-2 virus is primarily transmitted between people through respiratory droplets and contact routes [4-5], while recently, there has been considerable difference of opinion on airborne transmission [6-13].

Moreover, with a primary mode of transmission through respiration and aerosol transmission [14-15], there is great risk to healthcare workers who are exposed to infected patients through the use of procedures such as intubations, aerosolized medication, handling of human body fluids as well as through routine patient checks [16]. Also, in addition to the general ease of contagion of SARS-CoV-2, there is also evidence that the virus can remain active on inanimate surfaces for up to three days [17-18]. These dynamics of SARS-CoV-2 transmission are especially important in hospital setups. Through person-to-person transmission and social activity, it is apparent that so-called super-diffusion events in hospital settings are liable for continued outbreaks and clusters. Also, new results on the transmissibility of coronaviruses from contaminated surfaces in hospital settings are now emerging [19]. Furthermore, despite numerous measures to contain these infections and prevent contagions, many cases of SARS-CoV-2 infections acquired in hospitals have been reported [17, 20]. Because the severe acute respiratory syndrome coronavirus (SARS-CoV-2) is so contagious, together with the fact that the frequency of healthcare related transmission of SARS-CoV-2 is very high in some cases [21], there is a clear need to implement infection control practices robustly in hospital setups, especially in their intensive care units (ICU) where confirmed as well as suspected COVID 19 (SARS-CoV-2) patients are treated.

Also, although there are several cases where hospital acquired SARS-CoV-2 infections have been reported, the actual route of infection transmission is often largely in doubt [22-23]. This lack of knowledge has negative consequences for public health, risk management, the control of hospital-acquired infections, and in medico-legal aspects. For respiratory viruses, the existing data for indirect methods of transmission, including indirect contact transmission involving contaminated objects or surfaces, are mostly limited, but there is some evidence through the use of stochastic transmission models. Also, a review of the literature for influenza viruses suggests a possible mode of transmission through fomites [24-25]. The transfer of infectious viruses may readily occur once a fomite is contaminated. Fomites can be contaminated with virus by direct contact with body fluids, contact with SARS-CoV-2 contaminated hands, or respiratory droplets landing directly on surfaces [26-27]. However, direct experimental evidence of human infection of viral transmission via fomite can be very difficult to establish in the face of widespread community transmission. Therefore in this paper, we analysed different possible sources of indirect infections in various segments of hospitals that treat large numbers of suspected as well as confirmed COVID-19 patients. As a part of healthcare system, we have also included COVID 19 testing laboratories and disease diagnostic centres for evaluation in our study

## Methods

### Ethical statement

All authors do not have conflicts of interest. No experiment is conducted on animals or human subjects.

#### Hospital selection

Five large hospitals were selected from the private as well as public sectors for this study from the Aurangabad city in India which was one of the hotspot during second COVID 19 wave. Most of the hospitals selected are categorized as large as they have over 100 bed accommodation for confirmed COVID 19 patients. These hospitals have three sections viz. outpost patient department, COVID 19 ward, and COVID 19 intensive care units (ICU). Similarly, two health diagnostic centres and two COVID 19 testing laboratories were also selected for sample collection to analyze surface contamination. Additionally, twenty sewage water samples were also collected in the vicinity of healthcare units to assess environmental sources of contamination by SARS-CoV-2.

#### Sample collection

Samples were collected from various units as depicted in Figure 1. A sample is defined as a swab collected from all the probable places/objects/devices in the hospital setup where infection of SARS-CoV-2 virus is possible. Swabs were collected using a sterile, wet cotton swab (soaked in VTM). For sample collection, each swab was gently rubbed over the suspected spots multiple times and transferred to the VTM tube for preservation [28]. Sample tubes were properly labelled and stored at 4 0C in an insulated sample transport box and carried to the COVID 19 testing laboratory.

**Figure 1.**
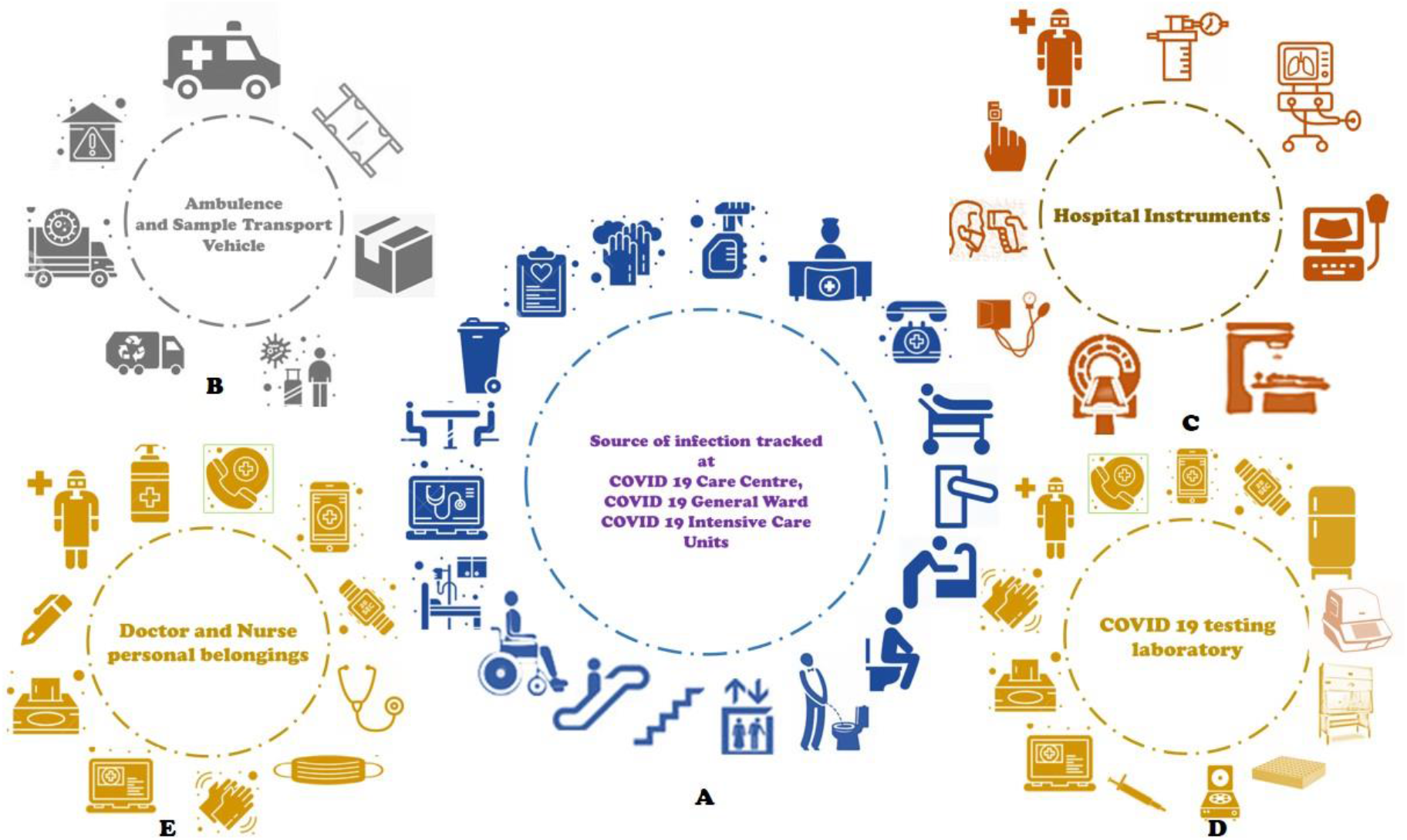
Swab samples collected from various segments of healthcare system for tracking SARC-Co-V-2 infection. A. Various hospital wards B. Ambulance and sample transport vehicles C. Hospital instruments D. COVID 19 testing laboratory E. Hospital staffers and their belongings

#### Sample processing

All collected samples were processed at the COVID 19 testing laboratory at the Paul Hebert Centre for DNA Barcoding and Biodiversity Studies, Aurangabad. Samples were processed following WHO laboratory guidelines [28]. For RNA isolation, MagRNA-II viral RNA extraction kits were used (Genes2Me Pvt. Ltd, Gurgaon, India) in a RNA purification machine (Thermo Fisher Flex, USA) following the manufacturers protocol. Isolated RNA was further tested using the QuantStudio Real-Time PCR system (Thermo Fisher Scientific Inc., USA) and the Meril RT PCR kit (Meril Diagnostics Pvt. Ltd., India) following standard operating protocols.

### Statistical analysis

A set of descriptive statistics of the laboratory testing was performed.

## Results

We assessed results from five hospitals, two diagnostic centres and two COVID 19 testing laboratories for SARS-CoV-2 infection during the study period. All processed samples qualified the quality control test specified for the detection of SARS-CoV-2 virus. A total of 558 samples were collected from various segments of the health care system (Figure 1). Of these, all 55 samples collected from ambulance and sample transport vehicles were found to be negative (Supporting material S1). Similarly, 64 samples collected from casualty wards were also found to be negative for SARS-CoV-2 (Supporting material S2).

Other segments of the hospital system tested for SARS-CoV-2 infection included the COVID 19 patient ward where we collected 125 samples. Of these, 9 (7.2%) samples were positive for SARS-CoV-2 infection (Table 1).

**Table 1.** Samples collected from COVID 19 patient ward at various hospitals

| Sr. No. | Source of infection tracked | Hospital surfaces tested for presence of SARS-CoV-2 Virus/Virus traces (Yes/No) |  |  |  |  |
| --- | --- | --- | --- | --- | --- | --- |
|  |  | H1 | H2 | H3 | H4 | H5 |
| 1. | Chair | No | - | No | No | - |
| 2. | Telephone Device | No | - | - | No | - |
| 3. | Doctors Apron | - | No | No | - | - |
| 4. | Refrigerator handle | - | - | No | - | - |
| 5. | Stethoscope | No | - | No | No | No |
| 8. | Air Vent | No | No | No | - | No |
| 9. | Bed-1 | No | No | No | No | No |
| 10. | Bed-2 | **Yes** | No | No | No | - |
| 11. | Bed-3 | No | Yes | No | No | No |
| 12. | Bed-4 | No | - | **Yes** | No | - |
| 13. | Bed-5 | No | - | No | **Yes** | - |
| 14. | Biomedical waste bin | No |  |  | No | No |
| 15. | BP Apparatus | - | - | No | No | No |
| 16. | Calculator | No | - | No | No | No |
| 17. | Curtains | - | - | No | - | - |
| 18. | Doctors cell phone | - | - | No | No | - |
| 19. | Files | No | No | No | No | **Yes** |
| 20. | Flooring | **Yes** | No | **Yes** | No | No |
| 21. | Floor Cleaning utensils | - | No | No | No | - |
| 22. | Hand Gloves | No | No | No | No | No |
| 23. | Glucometer | - | - | No | - | - |
| 24. | Elevator key board and railing | No | - | - | - | - |
| 25. | Mask-1 | No | - | No | - | No |
| 26. | Mask -2 | - | - | No | - | - |
| 27. | Medicine Trolley | No | No | No | No | - |
| 28. | Nebulizer | - | - | No | - | - |
| 29. | O<sub>2</sub> Mask |  |  |  |  | No |
| 30. | Oxygen cylinder | - | - | - | No | - |
| 31. | Pulse Oxymeter | No | - | No | No | No |
| 32. | Patient mobile | - | - | No | - | - |
| 33. | Patient File | No | No | No | - | No |
| 34. | Pen | No | No | **Yes** | No | No |
| 35. | PPE Kit | No | No | - | No | No |
| 36. | Punching Machine | No | - | - | - | No |
| 37. | Stapler | - | - | No | - | - |
| 38. | Stretcher | - | - | No | - | - |
| 39. | Electric Switches | No | No | No | No | No |
| 40. | Table | No | No | **Yes** | No | No |
| 41. | Thermometer | No | - | No | No | No |
| 42. | Toilet-1 | - | - | - | - | No |
| 43. | Toilet-2 | No | - | No | No | - |
| 44. | Wash Basin | No | - | No | No | - |
| 45. | Door Handle 1 | - | No | - | - | - |
| 46. | Door Handle 1 | - | No | - | - | - |
| 47. | Face Circle | - | - | - | - | - |

*H-Hospital*

In the ICU, out of 193 swabs collected, 9 (4.7%) samples were positive for SARS-CoV-2 (Table 2). At the outpost department of three hospitals, 20 samples were collected, and of these, one sample was found to be positive (Table 3).

**Table 2.** Samples collected from COVID 19 Intensive care unit at various hospitals

| Sr. No. | Source of infection tracked | Hospital surfaces tested for presence of SARS-CoV-2 Virus/Virus traces (Yes/No) |  |  |  |  |
| --- | --- | --- | --- | --- | --- | --- |
|  |  | H1 | H2 | H3 | H4 | H5 |
| 1. | Air Vent | - | - | No | - | No |
| 2. | Bathroom | - | No | - | No | - |
| 3. | patient File | No | No | - | No | No |
| 4. | Bed-1 | No | No | No | No | No |
| 5. | Bed-2 | No | No | No | No | No |
| 6. | Bed-3 | No | No | No | No | - |
| 7. | Bed-4 | - | No | No | - | - |
| 8. | Bed-5 | - | - | No | No | - |
| 9. | BP Apparatus | No | No | - | No | No |
| 10. | Chair | - | No | No | No | - |
| 11. | Cupboard | - | No | - | No | No |
| 12. | Curtain | No | No | No | No | No |
| 13. | Doctors Gloves | **Yes** | No | No | **Yes** | - |
| 14. | Door Handle | No | No | No | No | No |
| 15. | Doctor's Watch | - | - | No | - | No |
| 16. | Doctor's Mobile phone | - | No | - | No | No |
| 17. | Trash bean | No | No | No | No | No |
| 18. | ECG Machine | No | - | No | No | - |
| 19. | Face shield | - | - | No | No | - |
| 20. | Table Fan | - | - | No | No | No |
| 21. | Files | No | - | - | No | **Yes** |
| 22. | Floor | No | No | Yes | No | No |
| 23. | Floor cleaning mob | - | No | No | - | No |
| 24. | Freeze Handle | No | - | - | - | No |
| 25. | Glucometer | No | - | No | - | No |
| 26. | Wall side railing | No | No | - | - | - |
| 27. | Infusion pump | No | **Yes** | - | No | No |
| 28. | Stand | - | No | No | No | No |
| 29. | Elevator keyboard, railing | - | No | - | - | No |
| 30. | Mask | No | No | No | No | - |
| 31. | Medicine Trolley | No | No | - | No | No |
| 32. | Patient cell phone | No | - | No | - | No |
| 33. | Monitors | No | - | **Yes** | No | - |
| 34. | Sign board | No | No | - | - | - |
| 35. | News Paper | - | No | - | No | - |
| 36. | O<sub>2</sub> cylinder | - | - | No | No | No |
| 37. | O<sub>2</sub> regulator | - | No | No | No | - |
| 38. | Oxymeter | No | No | No | - | No |
| 39. | Patient file | - | No | No | - | No |
| 40. | Patient handkerchief | - | - | No | - | - |
| 41. | Patient monitor-1 | No | - | - | No | - |
| 42. | Patient monitor-2 | No | No | No | **Yes** | - |
| 43. | Patient monitor-3 | No | - | - | No | - |
| 44. | Patient monitor-4 | No | - | - | No | - |
| 45. | Patient O<sub>2</sub> Mask-1 | - | No | - | No | - |
| 46. | Pen | - | - | - | No | - |
| 47. | PPE Kit | No | No | No | No | - |
| 48. | Punching Machine | - | No | No | No | - |
| 49. | Sanitizer bottle | - | - | - | No | - |
| 50. | Scissor | - | - | No | - | - |
| 51. | Stapler | No | - | - | No | - |
| 52. | Sterile bottle | - | - | No | - | - |
| 53. | Stethoscope | No | No | - | - | No |
| 54. | Stretcher | No | No | - | No | - |
| 55. | Suction Machine | - | No | No | - | - |
| 56. | Electrical Switches | No | No | No | No | No |
| 57. | Inspection Table | - | No | No | - | No |
| 58. | Telephone | - | - | No | No | - |
| 59. | Thermometer | No | No | - | - | - |
| 60. | Ventilator-1 | No | - | No | No | No |
| 61. | Ventilator-2 | No | **Yes** | - | No | No |
| 62. | Ventilator-3 | - | No | No | - | No |
| 63. | Ventilator-4 | - | No | - | - | - |
| 64. | Wash Basin-1 | - | - | - | No | - |
| 65. | Wash Basin -2 | No | No | No | - | No |
| 66. | Water bottle | - | - | No | - | - |
| 67. | Water tank | - | - | - | - | **Yes** |
| 68. | Wheel chair | - | No | - | No | No |

**Table 3.** Samples collected from Out Post Department (OPD) at various hospitals

| Sr. No. | Source of infection tracked | Hospital surfaces tested for presence of SARS-CoV-2 Virus/Virus traces (Yes/No) |  |  |  |  |
| --- | --- | --- | --- | --- | --- | --- |
|  |  | H1 | H2 | H3 | H4 | H5 |
| 1. | Mouse 1 | - | - | - | - | - |
| 2. | Mouse 2 | - | - | - | - | - |
| 3. | Mouse 3 | - | - | - | - | - |
| 4. | Air Vent | No | - | - | - | - |
| 5. | Doctors Apron | No | - | - | - | - |
| 6. | Chair | No | - | - | - | - |
| 7. | Hand Railing | No | - | - | - | - |
| 8. | Keyboard | No | - | - | - | - |
| 9. | Pulse Oxymeter | No | - | - | - | - |
| 10. | Printer | No | - | - | - | - |
| 11. | Table | No | - | - | - | - |
| 12. | Oxygen Cylinder | - | No | - | - | - |
| 13. | Printer | - | - | - | - | - |
| 14. | Curtains in the Reception section | - | - | - | - | - |
| 15. | Reception: Patient bench | - | - | - | - | - |
| 16. | Reception: Table | - | - | - | - | - |
| 17. | Reception: Water tank | - | - | - | - | - |
| 18. | Sample Collector Gloves-1 | - | **Yes** | No | - | - |
| 19. | Sample Collector Gloves-2 | - | No | No | - | - |
| 20. | Sample Collector PPE Kit-1 | No | No | No | - | - |
| 21. | Sample Collector PPE Kit-2 | No | No | No | - | - |
| 22. | Security staff table | - | No | - | - | - |

*H-Hospital*

Another important segment of healthcare system during the current pandemic included the health diagnostic centres. We collected 38 swabs from two diagnostic centres, and of these, five samples were positive for SARS-CoV-2 infection accounting for 13.1% positivity (Table 4).

**Table 4.** Samples collected from health diagnostic centre

| Sr. No. | Source of infection tracked | Hospital surfaces tested for presence of SARS-CoV-2 Virus/Virus traces (Yes/No) |  |
| --- | --- | --- | --- |
|  |  | D1 | D2 |
| 1. | Inspection Table | No | - |
| 2. | Key Board 1 | - | **Yes** |
| 3. | Key Board 2 | - | No |
| 4. | keyboard 3 | - | No |
| 5. | Machine Keyboard right | - | **Yes** |
| 6. | Machine keyboard Left | - | No |
| 7. | Machine Stretcher | No | No |
| 8. | Monitors | No | - |
| 9. | Sonography Patient table | - | No |
| 10. | Sonography abdominal Probe | No | No |
| 11. | Sonography AC remote | - | No |
| 12. | Sonography curtain | No | No |
| 13. | Sonography door handle | No | No |
| 14. | Sonography door handle | - | No |
| 15. | Sonography Keyboard | - | No |
| 16. | Sonography Monitor | - | No |
| 17. | Sonography Napkins | - | No |
| 18. | Electric Switch 1 | No | No |
| 19. | Electric Switches 2 | No | No |
| 20. | Table | No | No |
| 21. | Toilet | No | No |
| 22. | Wash Basin | No | - |
| 23. | X-ray table | - | No |
| 24. | X-ray apron | - | **Yes** |
| 25. | X-ray fixer | **Yes** | No |
| 26. | X-ray machine | - | No |
| 27. | X-ray Screen | - | No |
| 28. | X-ray patient seating stool | **Yes** | No |

*D-Health diagnostic facility*

At the COVID 19 testing laboratories, out of 44 swabs collected, three were positive and 2 were inconclusive for SARS-CoV-2 infection (Table 5).

**Table 5.** Samples collected from COVID 19 testing laboratory and sample collection Facility

| Sr. No. | Source of infection tracked | Hospital surfaces tested for presence of SARS-CoV-2 Virus/Virus traces (Yes/No) |  |
| --- | --- | --- | --- |
|  |  | L1 | L2 |
| 1. | Sample Carrier Box (02) | No | No |
| 2. | Sample Carrier Box (02) | No | No |
| 3. | Sample Carrier (Belt) (02) | No | No |
| 4. | Sample Carrier (Belt) (02) | - | No |
| 5. | Record Register | - | No |
| 6. | Sample Receiving Surface/Floor | - | No |
| 7. | Door Handle (RNA isolation Room) | - | No |
| 8. | Centrifuge machine | - | No |
| 9. | Pipettes (05 nos.) (RNA isolation section) | Yes | No |
| 10. | Biosafety Cabinet (02) | - | No |
| 11. | Automation RNA purification machine | - | No |
| 12. | Chemical Bottles | No | No |
| 13. | Refrigerator | No | No |
| 14. | Handling Surface | No | No |
| 15. | Chairs RT PCR room | No | No |
| 16. | PPE Kit | No | No |
| 17. | Mask | - | No |
| 18. | Face Shield | - | No |
| 19. | Hand Gloves | - | No |
| 20. | Shoes Cover (04) | - | No |
| 21. | Downing/Doffing Area | - | No |
| 22. | RNA Plate (04) | - | **<u>IC</u>** |
| 23. | VTM Racks (04) | - | No |
| 24. | Record Books | - | No |
| 25. | Door Handle | - | No |
| 26. | Pipettes (Master Mix section) | - | No |
| 27. | Pipettes (RT PCR section) | - | No |
| 28. | Laminar Flow (Master Mix section) | - | No |
| 29. | Laminar Flow (RT PCR section) | - | No |
| 30. | Refrigerator | - | **<u>Yes</u>** |
| 31. | PCR Machines | - | **<u>Yes</u>** |
| 32. | Handling Surfaces (RT PCR section) | - | No |
| 33. | PC (RT PCR section) | - | **<u>IC</u>** |
| 34. | Chairs Data entry side | No | No |
| L-COVID 19 testing laboratory |  |  |  |

All samples collected from a mortuary were negative for SARS-CoV-2 (Supporting material S3). Finally, out of 20 sewage water samples collected in the vicinity of healthcare settlements, six were found to be positive for SARS-CoV-2 (table not shown).

## Discussion

The severe acute respiratory syndrome (SARS-CoV-2) virus is known to be very contagious [29] but still there is considerable debate on its mode of spread among human subjects [1, 3, 15, 30-31]. However, the journey of a person with an infection of SARS-CoV-2 may give some clues. This path usually starts with laboratory sample collection [32], then sample testing [33] (Drame et al., 2020), an OPD visit [34], and then transfer to a COVID 19 ward [35] and possibly further transfer to an ICU [36].

During the second wave of SARS-CoV-2, the ever growing number of patients moving along this path have put enormous pressures on hospital infrastructures. This has resulted in expanding hospital numbers and conversions of regular hospitals into COVID 19 care units and COVID 19 ICU wards [36]. In several instances, the number of beds in the same space increased, or normal hospital beds were converted to oxygen beds/ICU beds [37]. It is not hard to imagine how such practices might have a negative impact on the patient health as well as the health of hospital staff, possibly resulting in co-infections and superinfections with SARS-COV-2 [38]. This may be true even when COVID-19 lockdowns and social isolation measures may be reducing the spread of the infection [39], many healthcare workers, including managers and support staff [40-41], have to face consequences by working in high-risk environments [42].

Several hospital-based studies have been performed to study air-sampling for transmission of SARS-COV-2, but have not shown of significant evidence for infection [7-13]. Studies focused on air sampling in the hospital environments were based on air sampling [43] and related methodological protocol development [44]. However, most of these studies underestimated the possibilities of infections derived from other sources of contamination among health workers at hospitals, diagnostic laboratories and diagnostic centres [45].

Our present study is novel because for the first time, we tracked the possible impact of infective surfaces at various segments of healthcare system with SARS-CoV-2 that may potentially infect hospital staff. In our study, the hospitals selected were large with over 100 bed capacity to ensure more possibilities of cross infectivity either through aerosols or contaminated surfaces [46]. Swab samples were collected from all possible sources where patients could have physical contact as shown in Fig. 1 [47]. At all such places, items, including machines and belongings were sampled for SARS-CoV-2 virus using methods routinely used in collection of human samples [28].

A total of 558 swabs were collected for analysis from various segments within healthcare settings as shown in Fig. 1. Of these, 5.19 percent (N=29) samples were positive and 94.80 percent (N=527) were negative (2 were inconclusive) (Table 1-5; Supporting material S1-S3). Although the positivity rate among the surfaces tested here is low, persistence of viruses on the surfaces for a few days may promote transmission to healthcare workers or to other patients because they often share common spaces, and fomites can spread through touch contamination followed by self-inoculation of the mucous membranes [47].

In such instances, the infections we tracked were found positive for doctor’s belongings including pens, record files and tables as well as patient bed and ward floors in the COVID 19 ward (Table 1). In the ICU ward area, infection was found on doctors gloves, infusion pumps, ventilator monitors, ventilators, and drinking water dispensers (Table 2). In the outpost department, the gloves of the swab collection staff was found to be positive for SARS-CoV-2 (Table 3). Similarly, at the health diagnostic centre, the desktop computer key board which was used to register patient data was found to be infected. The key board associated with CT scan machine at one diagnostic centre was also positive for infection. Similarly, the drinking water facility and the X-ray technician’s apron was positive for SARS-CoV-2 (Table 4). At COVID 19 testing laboratories, the RT PCR machines and refrigerators used for storage of samples were positive although two samples collected from the surface of a 96 well plate used for carrying the isolated RNA samples and the RT PCR machine linked computer screen were inconclusive (Table 5). Finally, all of the samples collected from ambulances and the morgue were negative (Supporting material S1).

Although signals for SARS-CoV-2 virus infection were found on contaminated objects or surfaces that may lead to indirect contact transmission to hospital staff and patients it is, however, not always known whether the virus is viable when using the RT PCR method. To address this, it is of paramount importance to determine the infectivity of viruses detected because the mere presence of viral material on a surface does not by itself establish that they can be transmitted to and infect another person [47]. But, the possibility that these viruses can survive for prolonged periods of time on different surfaces suggests that they can serve as reservoirs for onward transmission from these surfaces to humans [48]. Additional studies on the persistence of coronaviruses outside of its host will be needed to clarify the role of contaminated surfaces on the transmission like those observed in cases of other SARS family viruses [49-50].

Finally, six samples of sewage water collected in the vicinity of the healthcare settlements were positive for SARS-CoV-2 infection, confirming results seen in other clinical studies [51-53] where prolonged faecal shedding of SARS-CoV-2 RNA was reported. However, to date there is no clinical evidence of water born SARS-CoV-2 viruses as a source of infection in humans [50, 54-56] nor are there currently any studies looking at the duration of viability of SARS-CoV-2 viruses in waste water [30, 57].

### Surface-to-person transmission modalities

The surface contamination we noticed in this study might have been transferred from the droplets deposit onto surfaces via gravitational sedimentation from an infected emitter [58]. Surfaces may also be contaminated via direct contact from the person to the surface. Contamination of surfaces via direct contact is very probable, especially for high-touch surfaces. So-called high-touch surfaces have been described by several studies [59-60]. Certain surfaces, particularly bed-parts such as bedrails, receive a large amount of contact both by the patient and the healthcare workers. In addition, touches to a surface generally do not occur in isolation, but rather as part of a sequence of touch events involving a variety of fomites. For example, when studying touch events of healthcare workers, a sequence of events could be defined as all of the touches a healthcare worker performs between their entry into a patient room and their exit [59]. During this time period, they may touch more than a dozen items in the room. Within these long events there may also be sub-events of touches that occur more frequently [59]. For example, touching a portable medical device and then the patient was the 5th most common sub-event. These findings demonstrate how interrelated all the items are in a room may be in terms of touch contacts. Given a certain probability to transfer a pathogen from a hand to a surface (as described in the transfer efficiency studies above), then it is possible that pathogens could be transferred to a surface and then to the patient or to another surface and then back again to the patient. If the fomite is on a piece of portable medical equipment, it may also be brought into multiple patient rooms where the sequence of touches involves touching the portable medical equipment and then the patient or a surface that the patient may later touch.

Although estimating the transfer of virus through a series of transfer events would be speculative at best, these studies reveal that transfer through a series of events is quite possible. Indeed, sequences of contact events where a fraction of pathogen is transferred with each successive touch as described above likely explain the findings by multiple studies that have reported widespread contamination of objects in the patient room, common work areas, and on portable medical equipment in the hospital after inoculating a few surfaces with cauliflower mosaic virus DNA markers [60-61].

Finally, our study has several advantages over previous studies as we covered all segments associated with health care systems. In this context we also offer recommendations for moving forward. Adherence to strict environmental cleaning policies is needed as routine hospital surface disinfection methods decrease viruses on the contaminated surfaces efficiently [62]. Also, several disinfectants are known to damage soft parts of hospital instruments like machine displays, computer screens, key boards, etc. In such cases, staff might be avoiding the use of these substances for disinfection because of a fear of damage. Therefore, to maximize the avoidance of indirect transmission of SARS-CoV-2 virus, suitable disinfectants must be recommended.

## Conclusion

The study results suggests that transmission of virus infection via contact with fomites from surfaces in a healthcare setup suggest is highly possible. Environmental contamination in rooms with COVID-19 patient has been widely observed because of repetitive contact cycles that can spread the virus from one surface to the other in healthcare settings. Based on the findings of this study, decontamination of surfaces that frequently are in touch contact throughout all segments of the healthcare system should constitute an important part of the infection control and prevention of COVID-19. We also recommend the selection of a non-reactive disinfectant for hospital monitors, devices, ventilators and computers so that active surface disinfection can be effected without damage to the devices.

## Data Availability

Relevant data is presented in the manuscript and i the supporting material

## Acknowledgements

Authors are thankful to the district administration for permitting to take swab samples from various hospitals. We are also thankful to COVID 19 testing laboratory staff at Paul Hebert Centre for DNA Barcoding and Biodiversity Studies for their efforts in sample testing. The language corrections and content editing done by Prof. David Haymer, authors are grateful to him.

