## Supporting material for "Infected surfaces as a source of transmissible material in healthcare settings dealing with COVID-19 patients"

**Supplementary material S2.** Samples collected from causality ward

| Sr. No. | Source of infection tracked | Hospital surfaces tested for presence of SARS-CoV-2 Virus/Virus traces (Yes/No) | | | | |
|---|---|---|---|---|---|---|
| | | H1 | H2 | H3 | H4 | H5 |
| 1. | BP apparatus | - | - | - | No | No |
| 2. | Bed 1 | - | - | - | No | No |
| 3. | Bed 2 | - | - | - | No | No |
| 4. | Bed 3 | - | - | - | No | No |
| 5. | Bed 4 | - | - | - | No | No |
| 6. | Bed 5 | - | - | - | No | No |
| 7. | CPU | - | - | - | No | No |
| 8. | Doctor’s mask | - | - | - | No | No |
| 9. | Doctor’s PPE 2 | - | - | - | No | No |
| 10. | Doctor’s PPE 1 | - | - | - | No | No |
| 11. | Door handle | - | - | - | No | No |
| 12. | ECG machine | - | - | - | No | No |
| 13. | Ward Flooring | - | - | - | No | No |
| 14. | Gloves (Nurse) | - | - | - | No | No |
| 15. | Keyboard | - | - | - | No | No |
| 16. | Mask (Nurse) | - | - | - | No | No |
| 17. | Medicine trolley | - | - | - | No | No |
| 18. | Monitor 1 | - | - | - | No | No |
| 19. | Monitor 2 | - | - | - | No | No |
| 20. | Mouse | - | - | - | No | No |
| 21. | Pulse Oxymeter | - | - | - | No | No |
| 22. | Oxygen cylinder | - | - | - | No | No |
| 23. | Patient Register | - | - | - | No | No |
| 24. | Stethoscope | - | - | - | No | No |
| 25. | Stretcher | - | - | - | No | No |
| 26. | Infusion Pump | - | - | - | No | No |
| 27. | Electric Switches | - | - | - | No | No |
| 28. | Inspection Table | - | - | - | No | No |
| 29. | Thermometer | - | - | - | No | No |
| 30. | Ventilator | - | - | - | No | No |
| 31. | Wash basin | - | - | - | No | No |
| 32. | Wheel chair | - | - | - | No | No |

H-Hospital

**Supplementary material S2.** Samples collected from causality ward

| Sr. No. | Source of infection tracked | Hospital surfaces tested for presence of SARS-CoV-2 Virus/Virus traces (Yes/No) | | | | |
|---|---|---|---|---|---|---|
| | | H1 | H2 | H3 | H4 | H5 |
| 1. | BP apparatus | - | - | - | No | No |
| 2. | Bed 1 | - | - | - | No | No |
| 3. | Bed 2 | - | - | - | No | No |
| 4. | Bed 3 | - | - | - | No | No |
| 5. | Bed 4 | - | - | - | No | No |
| 6. | Bed 5 | - | - | - | No | No |
| 7. | CPU | - | - | - | No | No |
| 8. | Doctor's mask | - | - | - | No | No |
| 9. | Doctor's PPE 2 | - | - | - | No | No |
| 10. | Doctor's PPE 1 | - | - | - | No | No |
| 11. | Door handle | - | - | - | No | No |
| 12. | ECG machine | - | - | - | No | No |
| 13. | Ward Flooring | - | - | - | No | No |
| 14. | Gloves (Nurse) | - | - | - | No | No |
| 15. | Keyboard | - | - | - | No | No |
| 16. | Mask (Nurse) | - | - | - | No | No |
| 17. | Medicine trolley | - | - | - | No | No |
| 18. | Monitor 1 | - | - | - | No | No |
| 19. | Monitor 2 | - | - | - | No | No |
| 20. | Mouse | - | - | - | No | No |
| 21. | Pulse Oxymeter | - | - | - | No | No |
| 22. | Oxygen cylinder | - | - | - | No | No |
| 23. | Patient Register | - | - | - | No | No |
| 24. | Stethoscope | - | - | - | No | No |
| 25. | Stretcher | - | - | - | No | No |
| 26. | Infusion Pump | - | - | - | No | No |
| 27. | Electric Switches | - | - | - | No | No |
| 28. | Inspection Table | - | - | - | No | No |
| 29. | Thermometer | - | - | - | No | No |
| 30. | Ventilator | - | - | - | No | No |
| 31. | Wash basin | - | - | - | No | No |
| 32. | Wheel chair | - | - | - | No | No |

H-Hospital

**Supplementary material S3.** Samples collected from the mortuary

| Sr. No. | Source of infection tracked | Hospital surfaces tested for presence of SARS-CoV-2 Virus/Virus traces (Yes/No) | | | | |
|---|---|---|---|---|---|---|
| | | H1 | H2 | H3 | H4 | H5 |
| 1. | Body cover (3 nos.) | - | - | - | No | - |
| 2. | Dead body cover (1 nos.) | - | - | - | No | - |
| 3. | Stretcher with dead body (2 nos.) | - | - | - | No | - |
| 4. | Patient apron | - | - | - | No | - |
| 5. | Doctor's mask | - | - | - | No | - |
| 6. | Doctor's gloves | - | - | - | No | - |
| 7. | Doctor's PPE | No | No | No | No | No |
| 8. | Flooring swab (section-I) | - | - | - | No | - |
| 9. | Flooring swab (section-II) | - | - | - | No | - |
| 10. | Empty Stretcher 1 | - | - | - | No | - |
| 11. | Empty Stretcher 2 | - | - | - | No | - |
| 12. | Empty Stretcher 3 | - | - | - | No | - |
| 13. | Room curtain | - | - | - | No | - |
| 14. | Electrical switch | - | - | - | No | - |
| 15. | Door handle | - | - | - | No | - |
